# Estimation of Annual Exposures and Antibody Kinetics Against Norovirus GII.4 Variants from English Serology Data, 2007-2012

**DOI:** 10.64898/2026.03.09.26347737

**Authors:** Kathleen M O’Reilly, James A Hay, Lisa Lindesmith, David Allen, Stephane Hue, Kari Debbink, Adam Kucharski, Ralph Baric, Judy Breuer, W John Edmunds

## Abstract

Norovirus in humans is highly contagious, causing diarrhoea and vomiting, and is especially common in young children. Winter incidence varies annually, and previous research indicates that the change of dominant norovirus variant is followed by high incidence, but having a clear mechanism to explain this observation could support better prediction of epidemics. Here we analyse unique norovirus serology blockade data from 656 children in England collected via opportunistic sampling between 2007-2012 using a mathematical model of multi-variant antibody kinetics to infer metrics such as annual attack rates and age-specific infection rates. Analysis reveals that overall infection rates were 204 infections per 1000 person-years (posterior median; 95% credible intervals: 188-221). Infection rates were lowest in children aged under 1 year at 164 infections per 1000 person-years (95% CrI: 121-209) and highest in children aged 5 years and older, at 252 infections per 1000 person-years (95% CrI: 212-288). The annual attack rate was highest in 2002, coincident with transition of the dominant variant to Farmington Hills, and high attack rates are frequently observed with emergence of new variants, but not always. Parameter estimates indicate moderate evidence for the immune imprinting hypothesis: a stronger antibody response to variants encountered earliest in life. Estimates of infection rates estimated here from serology are higher than incidence reported within similar settings based on disease only and is consistent with considerable asymptomatic infection. The combined use of multi-variant antibody data and a mathematical model provide key insights on the natural history of norovirus variants which can inform epidemic planning.

## Introduction

Norovirus is a highly contagious and common viral pathogen responsible for acute gastroenteritis worldwide. For most individuals, infection can be asymptomatic or result in intestinal disease symptoms such as vomiting and stomach cramps and typically resolves without requiring medical care. In vulnerable groups such as infants, the elderly or immunocompromised, norovirus infection can result in more severe disease and complications through nosocomial transmission and may require hospitalisation (1). While norovirus is the most common cause of gastrointestinal disease in the UK, norovirus-associated mortality within Europe has been estimated as 0.2 (95% uncertainty interval 0.1–0.2) deaths per 100,000 persons (2), which is at least 100-fold less than equivalent estimates for all-cause influenza-like illness (3) but remain a significant source of infection-induced mortality. Within the norovirus genus, the GII.4 genotype has accounted for most human norovirus outbreaks in recent decades. Variants within the GII.4 genotype are numerous and undergo natural variation that evolves and evades the host immune response, leading to recurring epidemics (4,5).

Large winter epidemics of GII.4 norovirus have been associated with the emergence of new variants. For example, vanBeek et al. report increased norovirus activity across Europe associated the emergence of the GII.4 Sydney variant (6), following its initial detection in Australia. Emergence of variants prior to 2012 also coincided with increased outbreaks, impacting hospitals, care homes and cruise ships (7–9). Understanding the mechanisms behind why variants emerge and continue to circulate is important as it could help predict future variant emergence events associated with epidemics and inform vaccine formulation. A curious aspect of norovirus epidemiology is that variants typically dominate circulation for a few years and are then replaced by a new variant. The exception to this observation has been the GII.4 Sydney (SY) 2012 variant which dominated in 2012 and has remained in circulation for over 10 years. Variants possess distinct genetic and antigenic features that allow them to evade pre-existing immunity within the population. However, studies have shown that previous exposure to GII.4 variants can confer variable levels of protection against subsequent GII.4 variants. This suggests the presence of cross-protective immune responses that target conserved regions of the virus or shared epitopes across different GII.4 variants.

Cross-immunity refers to the ability of the immune system, primed by exposure to one variant of a pathogen, to provide partial or complete protection against related variants. Cross-immunity is a common attribute of viral infections, and has previously been shown to drive evolution of viruses and modulate the immune response, best exemplified in the epidemiology of SARS-CoV2, influenza A and B (10,11). Classic measures of neutralising antibody responses are the preferred correlates of protection and require measurements of homotypic and cross-immunity, but are unavailable for norovirus. Instead, blockade assays based on preventing virus-like particle (VLP) engagement of receptor ligands have been developed by a few research groups (12–14), and have been shown to be an accurate correlate of immune protection. Studies using monoclonal antibodies specific to norovirus variants from mice illustrate strong blockade response against the same variant, and a limited response against different variants (15–18). Additionally, where convalescent human sera have been explored, a higher percentage of sera blocks heterotypic VLPs variants than homotypic VLPs variants which is especially apparent in adults (15). Infants with GII.4 Sydney 2012 gastroenteritis have the strongest blockade response to the Sydney 2012 variant (5). However, convalescent adults with recent GII.4 DenHaag-2009 or Sydney-2012 infection have been shown to have a stronger blockade response to previously circulating variants (5). These observations have led to the development the “immune imprinting” hypothesis (5,19,20), where individuals gain the strongest immune protection against variants encountered earliest in life. At a population level, the impact of cross immunity and immune imprinting on norovirus epidemiology is uncertain but can be explored using cross-sectional data and population models of infection.

In this study, we aim to understand the relationship between cross-immunity and epidemiological trends of norovirus from cross-sectional serology. We analyse this data with a statistical model which jointly estimates multi-strain antibody responses and individual-level norovirus infection histories. These estimates are used to infer annual and age-specific infection rates which will improve our understanding of norovirus epidemiology. Previous analysis of the data using simple catalytic models illustrated an increase in titre with age, but no discernible differences according to variant (21). We propose that use of a more complex model designed to capture individual infection histories will provide a more nuanced and informative analysis of the data.

## Results

### Description of data

A total of 656 serum samples from children aged between 8 months and 7 years of age at the time of sampling were available for analysis. The sera are from opportunistic samples derived from routine microbiological testing that are submitted voluntarily from hospital laboratories throughout England. Children were born between 2001 and 2011, while sera were collected between the norovirus seasons 2007 and 2012 (Appendix, Table A1). Data collection was cross-sectional, with 1 sample per child and IC_50_ blockade responses measured to GII.4 variants Farmington Hills (FH-2002), Den Haag (DH-2006), New Orleans (NO-2009) and Sydney (SY-2012). Antibody titres gradually increase as age increased but also revealed equal titres to some variants with little evidence of circulation up until the point of sampling (Appendix, Fig. A1). In 2009 when the NO-2009 variant emerged, a higher titre is observed across all age-groups when compared to the remaining biomarkers. There is limited opportunity for direct observation of serological responses following exposure to the DH-2006 variant as the first sample year is 2007, and there were comparatively fewer sera sampled in 2012 limiting inference of exposures to SY-2012 (n=42 where other years had over 100 samples). The “age of child at first variant isolation” was calculated for each variant and the IC_50_ responses plotted to examine visual evidence for immune imprinting. The IC_50_ response was highest when each variant had emerged near the childs’ birth year: this was most apparent for DH-2006 and NO-2009 where the data availability best aligns with emergence of these variants (Fig. 1).

**Figure 1.**
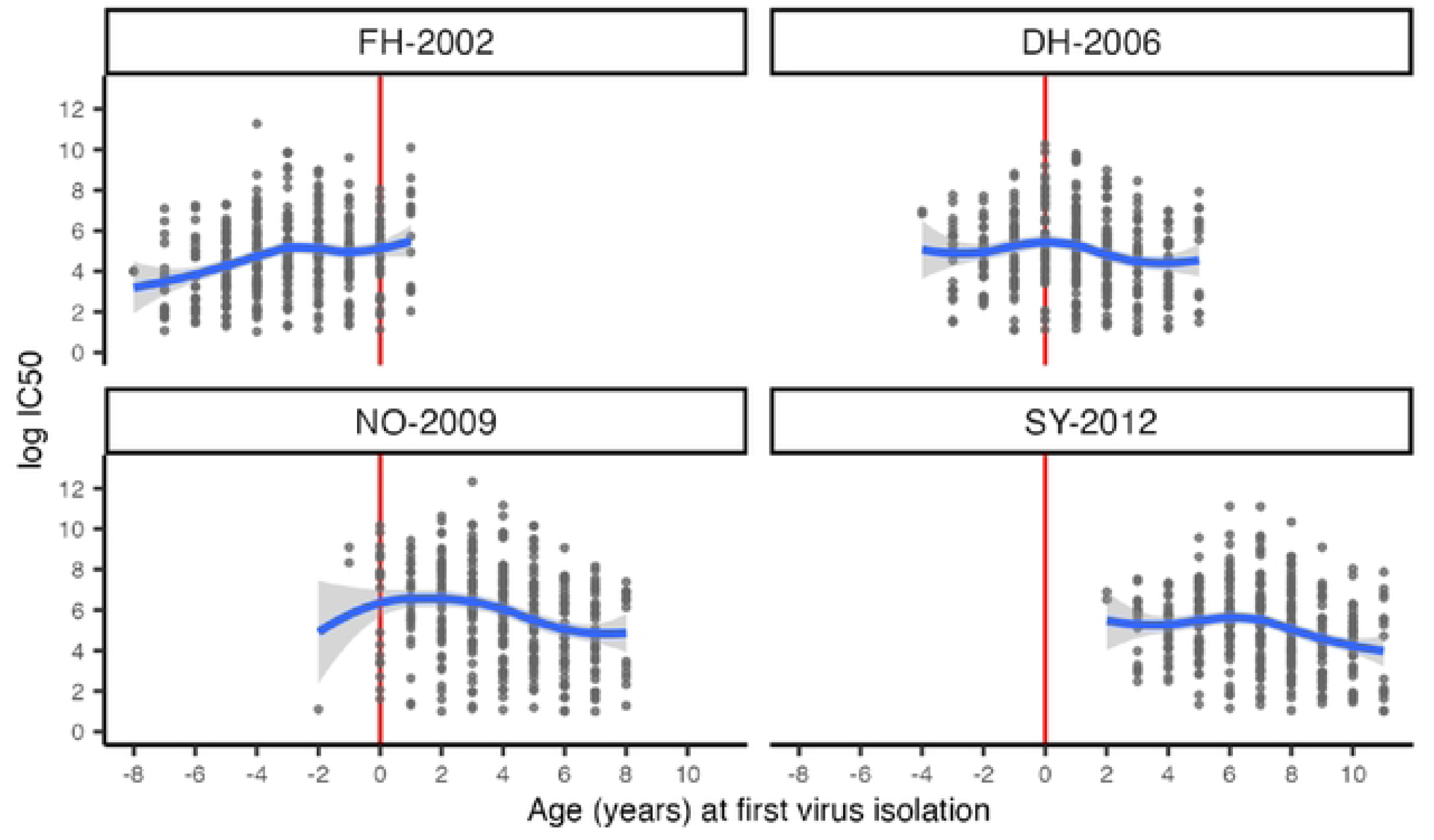
Relationship between log IC50 titre and age at time norovirus strain was first isolated. Vertical red line depicts individuals born when the strain first began circulating. Each dot shows one observation. Blue line and grey ribbon show fitted LOESS smoothing spline and 95% confidence intervals.

### Norovirus variant dynamics and antibody cross-reactivity

Between 2003 and 2012 there were five norovirus GII.4 variants that dominated reported cases of norovirus in surveillance data within England (Fig. 2, see materials and methods for details on data). The dominant variant switched in the winter of 2004 (FH-2002 to Hunter 2004), 2006 (Hunter to DH-2006), 2009 (DH-2006 to NO-2009), and 2012 (NO-2009 to SY-2012), with some evidence of earlier circulation for several variants (eg. DH-2009 and SY-2012) and emergence of other variants which did not dominate (eg. Yerseke and Apeldoorn).

**Figure 2.**
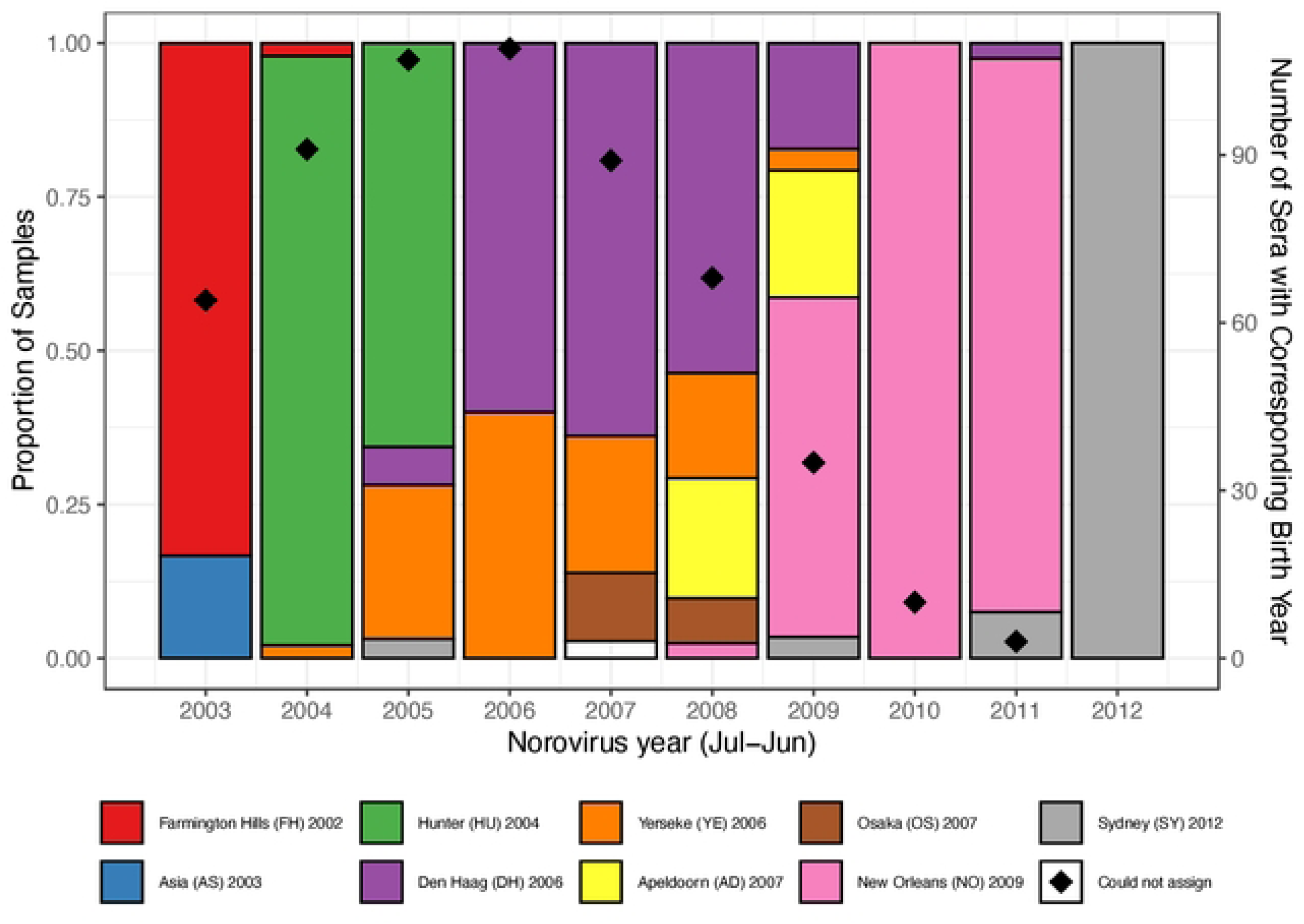
Gll.4 human norovirus variants detected through surveillance in England. Surveillance consists of **testing a** sample of confirmed **symptomatic** norovirus **cases** submitted **to UKHSA** to **identify** the **norovirus variant.** Coloured **bars** indicate the Gll.4 **variant** identified **by norovirus year** (July-June). **Black diamonds** indicate the number **of sera used** in the **analysis with** the corresponding **year** being the first **year** of **life,** consequently the dominant variant for that year was the likely first infection. A total of 74 individuals and their corresponding sera were born prior to July 2003 and are omitted from the plot.

To understand the antigenic relationships between these variants, sera from mice immunised with a specific variant VLP were tested for their antigenic response to all other variants and analysed using antigenic cartography. Two challenge studies are available for norovirus; a study of five variants that uses the same blockade as our serology data (‘Debbink data’ (18)), and another that includes eight variants but uses a different blockade (‘Kendra data’ (16)). Antigenic cartography (22) makes use of multidimensional scaling to reduce the complex relationships of individual sera against multiple antigens down to a 2-dimensional relationship, where the Euclidean distance between two antigens on the map is proportional to their antigenic relatedness (see materials and methods for full details). Within each publication antigenic maps are provided, but for consistency the raw data are analysed to reproduce the maps here. The results from each dataset were centred on the FH-2002 variant as a reference for comparison (Fig. 3A and 3B).

**Figure 3.**
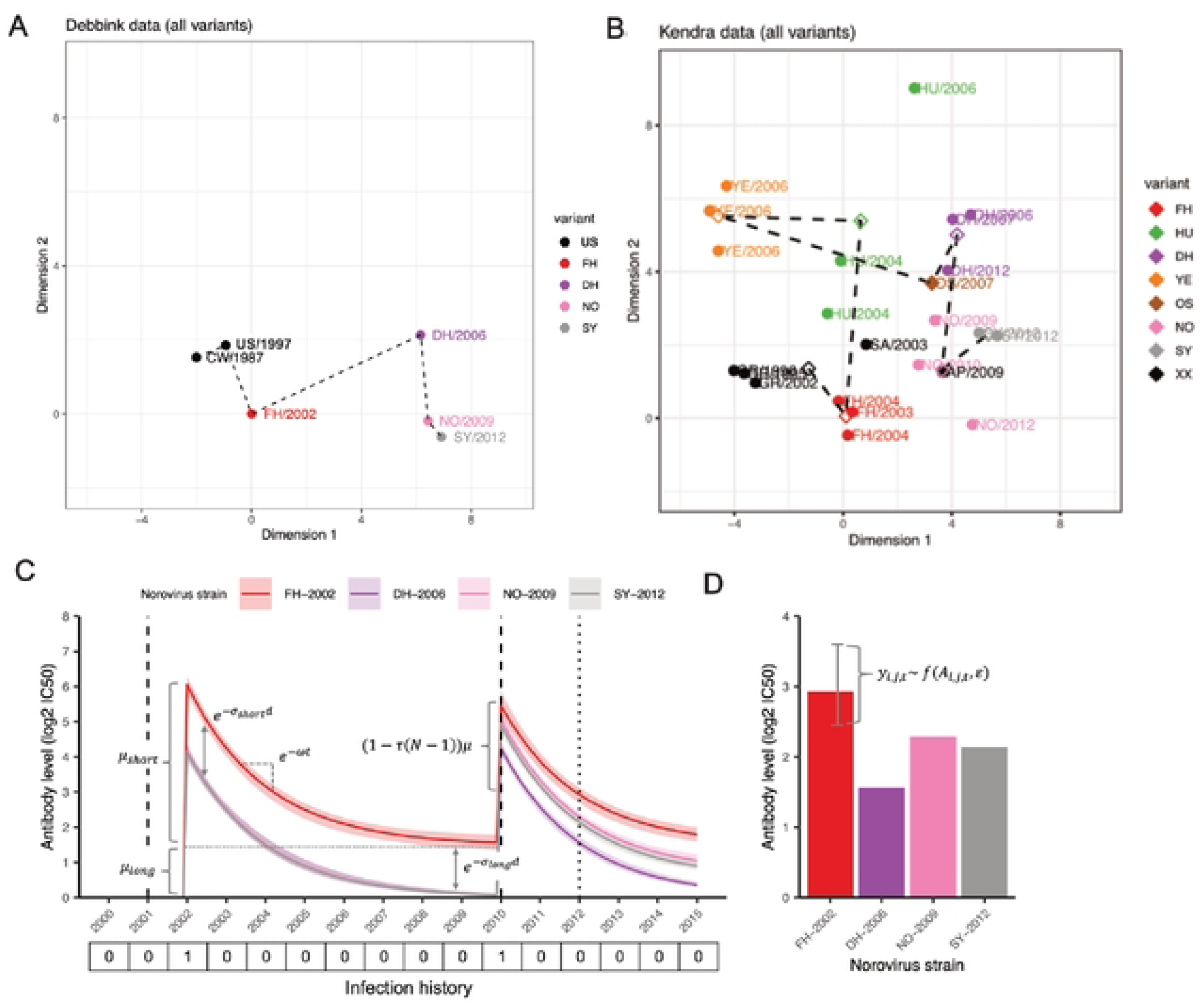
Antigenic cartography of norovirus Gll.4 variants and antibody kinetic models used in the model framework. (**A and B**) The estimated antigenic relationships are centred on the common variant Farmington Hills (FH) and temporal changes indicated by the dashed lines. Each variant is coloured, indicating common variants across datasets (FH, Den Haag (DH), New Orleans (NO), Sydney (SY)). (C) Illustration of model structure using posterior estimates (posterior median and 95% Crl) from fitting to IC50 data. Schematic shows a hypothetical infection history of an individual infected in 2002 and 2010, resulting in antibody kinetics to each of the four measured norovirus strains over time. Vertical dashed lines show time of infection, matching the infection history vector below. Vertical dotted line shows timing of serum sample collection. Inset text depicts model parameters including long-term and acute boosting (*µ_long_* and *µ_short_*); the acute boost waning rate(*ω*); the cross-reactivity gradient as a function of antigenic distance, *d,* from the long-term and acute boost (*σ_long_* and *σ_short_*); and the degree of boosting suppression from each successive boost (τ). (D) Simulated set of observations from a single serum sample taken in 2012 against each of the four norovirus strains. Bars show posterior median predicted titre for individual *i* at time *t* =2012 against each strain *j, A_i,j,t_,* Error bars show hypothetical range of observations under the observation model, *f*(*A_i,j,t_*, ε).

The Debbink map consists of data from mice infected with variants United States (US-1997), FH-2002, DH-2006, NO-2009, and SY-2012 (Fig. 3A). The antigenic distance between FH-2002 and DH-2006 is largest, followed by NO-2009, and the distance between NO-2009 and SY-2012 is comparatively small. These variants were commonly detected from 2003 to 2012 in England, however challenge data for Hunter (HU-2004) was not available in the Debbink data.

Within the Kendra data the antigenic responses to the HU-2006 variant are available, although the three antigens that correspond to HU-2004 cover a large dimensional space, resulting in some uncertainty in placement (Fig. 3B). The HU-2004 variant may no longer have been circulating by the time that children in this study are born, potentially limiting the impact of this uncertainty. There is some uncertainty on the placement of NO-2009 between datasets; the Debbink data suggests some similarity between NO-2009 and SY-2012, while the Kendra map has multiple isolates of the DH-2006, NO-2009 and SY-2012, resulting in some uncertainty in relatedness. The source of these uncertainties are due to use of different antigens and assays within each dataset, and are not possible to resolve. Instead, we report results using the Debbink map as the main estimates but performed sensitivity analyses using the Kendra map.

### Description of mathematical model

Sera from a total of 656 children aged 1-7 years at the time of sampling were examined for their exposure to norovirus GII.4 variants FH-2002, DH-2006, NO-2009 and SY-2012 using the serosolver R package. Serosolver (23) uses cross-sectional or longitudinal serological data in combination with an antigenic map to estimate individual-level infection histories from birth, age– and year-specific virus attack rates, and parameters describing homotypic and heterotypic (ie. cross-reactive) antibody responses. Each infection is assumed to generate an antibody response consisting of both a long– and short-term response to each variant, where we assume the short-term response wanes exponentially over time (Fig. 3C). The homotypic response is assumed to be the strongest, and heterotypic responses are assumed to decrease relative to the homotypic response proportional to the antigenic distance between variants. Repeated homotypic and heterotypic responses form an individual’s antibody landscape against all variants; serosolver uses a Bayesian framework to estimate infection histories and antibody kinetics parameters consistent with the observed data (24). Further, we test for evidence of immune imprinting by allowing for a declining homotypic antibody response with each successive infection. However, it should be noted that a declining homotypic antibody response with multiple infections may also correspond to other mechanisms of biomarker kinetics such as immune senescence or an antibody ceiling effect.

### Population-level incidence trends inferred from serosolver

Under this model, we estimate the overall norovirus infection rate to be 204 infections per 1000 per year (posterior median; 95% credible intervals: 187-220). Infection rates were lowest in 0-1 year olds at 163 infections per 1000 per year (95% CrI: 120-211) and increased with age up to 251 infections per 1000 per year (95% CrI: 210-289) in the 5+ age group (Table A2). Annual attack rates were estimated to vary considerably between norovirus years (Fig. 4A). The highest attack rate during the study period was in 2002 with an attack rate of 0.43 infections per year, although the confidence intervals were large (95% CrI: 0.30-0.55). The attack rate in 2009 was also high, being 0.33 infections per year (95% CrI: 0.29-0.27), and variant replacement was observed in both years. In contrast the 2006 season, where transition from FH-2002 to DH-2006 was reported, had a very low estimated attack rate at 0.04 (95% CrI: 0.01-0.16), highlighting the temporal variation in norovirus season intensity and that not all years with variant replacement are coincident with high attack rates. Reweighting the age distribution of the sample to match the true age distribution in the population based on census data did not have a major impact on the estimated attack rates (Appendix, Fig. A2). Age-year attack rates suggested a higher attack rate across most years for children less than 1 years of age, and broadly similar attack rates within each age group for older children (Appendix, Fig. A3), although it should be noted that these estimates are based on a smaller sample of children.

**Figure 4.**
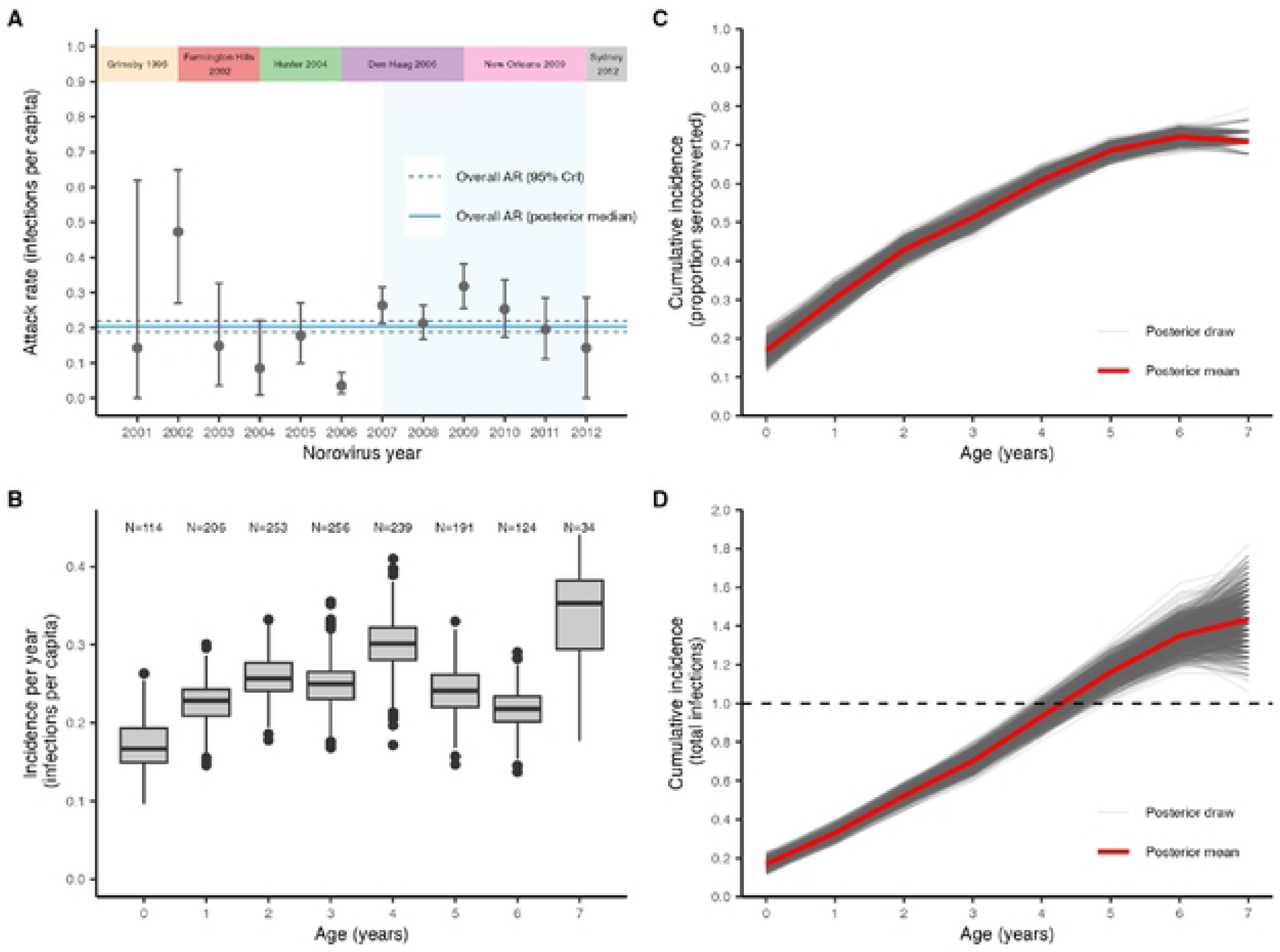
(**A**) Adjusted annual attack rates. Points and error bars show the posterior median and 95% credible intervals respectively. The horizontal blue lines show the overall adjusted posterior median attack rate across all time periods: 204 (posterior median; 95% Crl: 187-220) per 1000 adjusted (200 (190–209) per 1000 people unadjusted). The dominant strain in each year is shown at the top. The blue shaded rectangle depicts the sample collection period. **(B)** Proportion of individuals infected per year of age. Box plots show first, second and third quartiles. (C) Cumulative proportion of individuals infected at least once by age. Grey lines show posterior draws. Red line shows posterior mean estimate. **(D)** Similar to (C) but showing the cumulative total number of infections over age.

### Timing and recurrence of norovirus infection in childhood

We estimated that infections occurred consistently throughout childhood, with 50% of children having experienced at least one infection by age 3 (Fig 4C). These infections are not uniformly distributed across the population. After increasing consistently with age up until 5 years, the percentage seroconverted plateaued at 68.5% (95% CrI: 65.6-70.9%) in the 5-year-old age group and then increased only marginally to 72.2% (95% CrI: 69.7-74.5%) in the 6-year-old age group. In contrast, the mean number of cumulative infections continued to increase with age (Fig 4D). This pattern is consistent with around 30% of the cohort being persistently immune to infection, or at least not demonstrating any detectable serological response; individuals inherently susceptible to infection continue to experience reinfections whereas inherently immune individuals never experience a detectable serological response. Broadly similar patterns in the cumulative number of infections can be seen when stratified by year of birth (Appendix, Fig. A4).

### Homologous and cross-reactive antibody responses to norovirus infection

Parameter estimates of antibody kinetics suggest a relatively small, sustained long-term homotypic response with moderate evidence of immune imprinting that reduces the magnitude of boosting from each successive infection (Table 1; Fig A2). The model also estimates a large, transient response which wanes to 50% after approximately 1.4 years, and to 95% after 6 years. Parameter estimates suggest considerable cross reactivity between variants from the short-term response, as indicated by the σ_*short*_ estimates approaching zero, and a more antigenically narrow long-term response indicated by a higher value for σ_*long*_ (Table 1; a lower value for σ corresponds to a greater range over antigenic space), further supporting the broad serological response to GII.4 infections. Estimates of the proportional decrease in boost per successive infection, which can be interpreted as a measure of immune imprinting, showed a moderate impact where the upper 95% credible interval estimates were approximately 10% when the exponential model for antibody waning was used. Fig. A5 (Appendix) shows how an individual’s antibody landscape is estimated to develop through early life given repeated infections.

**Table 1.** Parameter estimates from the Norovirus serology data collected in England, 2007-2012. Parameter estimates are provided while using antigenic cartography from both the Debbink and Kendra datasets. Parameters are estimated using only IC_50_ data.

| Symbol | Meaning | Parameter estimate exponential model- Debbink | Parameter estimate exponential model – Kendra | Parameter estimate linear model- Debbink | Parameter estimate linear model – Kendra |
| --- | --- | --- | --- | --- | --- |
| $\mu_{long}$ | Long-term antibody boosting | 1.51 (1.26, 1.73) | 1.88 (1.58, 2.23) | 3.17 (2.98, 3.37) | 3.37 (1.74, 3.95) |
| $\mu_{short}$ | Short-term antibody boosting | 4.5 (4.24, 4.75) | 3.97 (3.55, 4.34) | 2.49 (2.2, 2.83) | 2.71 (2.14, 4.04) |
| $\sigma_{long}$ | Cross-reaction (retained) | 1.56 (1.06, 1.96) | 0.427 (0.281, 0.613) | 0.0672 (0.0582, 0.0751) | 0.143 (0.107, 0.273) |
| $\sigma_{short}$ | Cross reaction (acute) | 0.0122 (0.00152, 0.0227) | 0.00523 (0.00017, 0.0193) | 0.0105 (0.000238, 0.038) | 0.00703 (0.000271, 0.0265) |
| $\tau$ | Immune imprinting, ie. proportion decrease in boost per successive infection | 0.0433 (0.00217, 0.0962) | 0.0521 (0.00578, 0.106) | 0.225 (0.171, 0.283) | 0.207 (0.165, 0.249) |
| $\omega$ | Acute waning | 0.511 (0.45, 0.573) | 0.595 (0.497, 0.742) | 0.485 (0.334, 0.595) | 0.464 (0.107, 0.639) |
| $\varepsilon$ | Standard deviation of the observation error distribution | 1.12 (1.07, 1.17) | 1.12 (1.07, 1.17) | 1.13 (1.09, 1.18) | 1.09 (1.04, 1.16) |

### Sensitivity analyses

To test the robustness of our model estimates to the choice of antigenic map and antibody kinetics model structure, we ran several sensitivity analyses varying these inputs to serosolver. We found that the choice of antigenic map (using data from the Debbink or Kendra study) made very little difference to both antibody kinetics parameter estimates (Table 1) and epidemiological trends (Appendix Fig. A6), with the exception of the long-term cross-reactivity parameter, σ_*long*_, which was estimated to be higher using the Debbink map than the Kendra map under the exponential waning kinetics model, and vice versa under the linear waning kinetics model. However, these differences did not lead to qualitatively different predictions.

In contrast, the antibody kinetics parameter estimates showed greater sensitivity to the choice of model. Although the choice of model did not affect inferred infection trends there were notable differences in the antibody kinetics model parameters. First, the exponential waning model led to a large transient response and smaller persistent response, whereas the linear waning model led to a larger persistent response and smaller transient response (Table 1). Furthermore, whereas both models did show some evidence of immune imprinting, the magnitude of the immune imprinting parameter was much greater under the linear waning model. However, the exponential model had an improved fit to the data compared to the linear model (illustrated by comparing the predicted antibody levels to the observed antibody levels, Appendix Fig. A7 and A8) so we have more confidence in these results.

## Discussion

Norovirus infection persists in the community, especially in young children because of the availability of susceptible individuals that is mediated through births, a variable and changing immune landscape and virus escape. We find that the response to initial norovirus infection is both variant specific and elicits protection across variants, enabling a gradual rather than acute build-up of immunity; as variants emerge the immune landscape facilitates persistence over several years. The unique serology data analysed here points towards the roles of repeated infections and an attack rate that varies by year. This finding can at least in-part explain why norovirus is a common gastrointestinal pathogen: GII.4 norovirus infections occur approximately every 4-5 years in children due to changing variants and a moderately reduced immune response to subsequent variants experienced by an individual.

The estimated incidence was highest in children aged 4 or 5 years, being approximately 250 infections per 1000 person years. This rate of infection is higher than other studies that typically report symptomatic disease but remains within expectations that include asymptomatic infections. A Dutch community cohort study of gastrointestinal illness in children reported an incidence of 312 norovirus cases per 1000 person years (25), broadly similar to our estimates. A UK community cohort from 2008 reported an incidence of 142.6 (95% CI 99.8-203.9) symptomatic norovirus cases per 1000 person years within the 0-4 years age group (26). Within the UK study the average disease incidence was higher in infants (although a small sample size meant large uncertainty in estimates): combining results from this and the UK (24) study suggests a higher probability of being symptomatic during primary norovirus infection. A recent cohort study exploring norovirus infection in young (<2 years) children in Australia (27) has also indicated a higher (39.2%) symptomatic rate than typically found. Infection rates likely peak at 4 or 5 years of age due to increased contact, largely through increasing school attendance and growing social networks (28), older ages may have similar contact rates but retain some immunity from previous norovirus infection thus limiting infection rates. While data from young children can be logistically challenging due to ethical implications and challenges with collecting sufficient volume of sera from young children, sera from older ages would be more challenging to interpret as many individuals would already have a broad antibody response to several norovirus genotypes and variants.

We also found that approximately 30% of children in our study had no serological response to infection despite the high attack rates estimated. This observation is consistent with previous evidence that between 20-30% of the UK population are resistant to norovirus GII.4 infection due to the nonsecretor status of human histoblood group antigen carbohydrates (29). This study estimated high attack rates coincident with emergence of FH-2002 and NO-2009, but a lower attack rate when DH-2006 emerged. The attack rates are consistent with UKHSA annual reports of norovirus from the 2000-2012 period (30). A low attack rate in 2006 could be influenced by antigenic similarity of the DH-2006 variant to the HU-2004 variant that was first detected in England in 2004, lessening the attack rate for DH-2006. However, the antigenic distance between HU-2004 and DH-2006 is not so different to other variants. Additionally, between 2005 and 2008 several norovirus variants are co-circulating, perhaps indicating low reproduction numbers of the circulating variants or a transitional phase in norovirus epidemiology that may have influenced the low attack rate.

Evidence for norovirus immune imprinting has now been identified in experimental studies (31), small datasets from convalescent sera (5,32) and cross-sectional serological data that incorporates a mathematical model (21). Immune imprinting has also been observed in influenza (19,33) and SARS-CoV-2 infection (34), with the addition of increasing influence of an individual’s age on the expected response to a variant. The main effect of immune imprinting on norovirus epidemiology is the increased potential for persistence. The immune response against norovirus will be strongest against the variant first encountered with subsequent responses to variants consisting of both homotypic and heterotypic responses, resulting in lower immune protection against current variants for most of the population. For vaccine development, these findings suggest that a strong immune response may be best achieved by vaccinating young children. The implications of this on identifying target age groups for vaccination is beyond the scope of this analysis, but previously published mathematical models for norovirus transmission (35,36) could be updated to explore the effects of age-specific vaccine efficacy. This would support development of the multi-valent norovirus vaccines in development (37,38).

This study considers only the norovirus genotype GII.4, which is within the diverse GII genogroup, and distinct from five additional genogroups (39). The GII.4 genotype comprises the majority of reported cases of clinical norovirus from infectious disease surveillance (40), and where epidemiological studies have been carried out, have also been the majority of human infections. A multi-genotype mathematical model fitted to English data suggests that the GII.4 genotype is more transmissible than other genotypes such as GI.3 (36). More recently in the UK and other high-income countries, emergence of an additional norovirus genotype (GII.17) has been reported (41), sometimes cocirculating with other dominant noroviruses. The reasons for this emergence remain unclear, and as this emergent genotype is of another genotype, is beyond the scope of this current analysis. The antigenic relationships between norovirus genogroups are not well defined, although microarray data (42) suggest considerable cross reactivity between genogroups, although it is based on antibody binding rather than antibody blocking and could indicate cross-reactivity. Further research, and especially interdisciplinary research between virologists and epidemiologists would likely improve our understanding of norovirus infection and immunity.

A weakness of this analysis is that the sera were collected just prior to emergence of the SY-2012 variant, where only a small proportion of individuals will have experienced SY-2012 infection. Although the sera were tested against the SY-2012 blockade, the response is likely associated with a combination of highly variable cross reactive antibody blockade patterns and low-level homotypic exposures. There were few sera available in 2012, resulting in wide confidence intervals for the attack rate. The study was designed to sample evenly across years of collection and the variants studied were those that were circulating at the time. Considering the potential importance the first viral infection, future data collection should match sera and variants examined to those in circulation from the time of birth to sample collection. While the analyses would have benefitted from exploration of additional variants, sera volume, typically low in young children, was a limiting factor. Additionally, it was not possible to estimate variant-specific antibody kinetic parameters which may have provided some insight on reasons for the extended period of SY-2012 circulation, largely due to the limited period that individuals were exposed to this virus.

The modelling analyses have several caveats and limitations. First, we did not model co-circulation of variants, assuming that only one antigenic variant was circulating in each year. Infectious disease surveillance largely supports a single dominant variant each year, but this assumption may miss infections with non-dominant variants and thus misattribute boosting to the wrong variant. Second, we tested two antibody kinetics models, one assuming exponential waning on the log scale over time and with antigenic distance, and one assuming linear waning. Both models generated reasonable fits and almost identical epidemiological trend estimates, but qualitatively different antibody kinetics predictions. While we focus on the exponential waning model due to its better fit to the data, we cannot conclude that one model is closer to the true model than the other, as there is limited longitudinal information in this cross-sectional data. For identifiability reasons, we also assumed that all serological responses were governed by the same parameter values, rather than assuming random effects on the antibody kinetics parameters. This simplification ignores true immunological variation in the serological responses to infection. Finally, we did not have external data with which to validate our inferred infections (e.g., virologically confirmed cases), and thus we could only evaluate our model performance based on its fit to the data rather than out-of-sample predictive accuracy.

In conclusion, application of a mathematical model to unique serology data for norovirus has illustrated the role of primary infection in shaping the immune response, and the variation in annual incidence. Modelling suggests a high attack rate often in years with newly emerging variants. The gradual build-up of broadly neutralising antibody creates population immunity to circulating variants, limiting the frequency and severity of new infections. Further, our analysis suggests a moderate reduction in an individual’s response to subsequent infections that is consistent with the immune imprinting hypotheses. This enables repeat infections and contributes to the persistence of norovirus in the community.

## Materials and Methods

### Infectious disease surveillance data

Surveillance data for norovirus in England is carried out by the Enteric Virus Unit, UK Health Security Agency (and equivalent prior organisations). Stool are collected from a sample of patients thought to be part of norovirus outbreaks and are referred to the Virus Reference Laboratory as part of sentinel surveillance. Suspected norovirus outbreaks are defined locally if there are two or more cases with episodes of vomiting (with a suspected infectious cause) or loose stool in a 24 hours period, or both. If norovirus is confirmed at a regional laboratory, a sample is submitted to the Reference Laboratory for genotype and variant characterisation. The genotype is identified using real-time PCR and genotype specific primers, and the variant is identified through analysis of the P2 domain via sequencing (43). The year of outbreak is described by norovirus season, which runs from the 1^st^ July each calendar year, ie. for 2003 the year runs from 1^st^ July 2003 to 31^st^ June 2004.

### Antigenic Cartography

Antigenic cartography is a method for representing the antigenic relationship between variants of antigenically variable pathogens (22). For pathogens with considerable genetic variation, each mutation may result in a change in amino acid composition, potentially resulting in a change in binding properties when interacting with the host. It is assumed that antisera to specific variants (or strains in other contexts) represent the reactivity against specific antigens: higher responses correspond to a specific variant response. This specific response may also result in some cross-reactivity between related variants, and it is this relationship that helps identify the antigenic similarity between variants. Antisera for multiple variants from primary infection with one variant (typically from challenge studies) provides information on the antigenic relatedness of the variants. This multidimensional data can be reduced down to 2 (or more) dimensions using multi-dimensional scaling. In this application we used the online tool “ACMacs” (https://acmacs-web.antigenic-cartography.org/) to provide estimated antigenic distances for variants present in the Debbink and Kendra datasets (15,16). The optimiser was run for 1,000 iterations and repeated three times to assess the reliability of the antigenic distances (no discernible differences were identified per run). The antigenic distances were outputted as a.csv file, plotted in R (version 4.5.0), and used as input parameters for serosolver.

### Serological Data

Sera from a total of 656 children aged between 1-7 years were collected within England between 2008-2012. The sera used in the analysis are publicly available, and their use was approved by the National Health Service Research Ethics Committee (ref. 17/EE/0269), London School of Hygiene and Tropical Medicine (reference LEO12196), and the University of North at Carolina Chapel Hill (18–0214). The sera are from the Public Health England Seroepidemiology Unit (PHE SEU), which is an opportunistic collection of residual serum samples from routine microbiological testing, submitted voluntarily each year from hospital laboratories throughout England. No formal consent was obtained from the parent/guardian as these sera are collected as part of routine microbiological testing. Further details are provided in Lindesmith et al (21). These sera were subjected to a surrogate neutralisation assay that correlates with an *in vitro* virus neutralisation assay and is a proposed correlate of protection (31,44–46). Both IC50 and avidity (measured as the slope of the Ab of VLP binding to ligand) were reported for the GII.4 norovirus variants FH-2002, DH-2006, NO-2009, and SY-2012. The serology data were log_2_ transformed to ensure normality of the data. Data are available https://zenodo.org/records/7547170 (as part of a previous publication).

### Serology Kinetics and Infection History Model (serosolver)

The serosolver method finds the combination of variants which an individual is most likely to have been infected with conditional on their serology profile, accounting for cross-reactive, transient antibody boosting and immune imprinting arising from repeated exposures to antigenically related variants.

#### Infection History Model

More precisely, serosolver is a Bayesian, hierarchical model which estimates the joint posterior distribution of individual infection histories, represented as vectors of latent binary variables, ***Z***_*i*_, and antibody kinetics parameters, θ, conditional on individual-level antibody profile data (23). ***Z***_*i*_ = [*Z*_*i*,1_,*Z*_*i*,2_,… *Z*_*i*,*j*_] where *Z*_*i*,*j*_ = 1 represents infection of individual *i* in norovirus season *j* with the dominant variant assumed to be circulating at that time and *Z*_*i*,*j*_ = 0 represents no infection. Individual infections were assumed to be Bernoulli variables distributed according to independent Beta-Bernoulli distributions for each time period and age group:

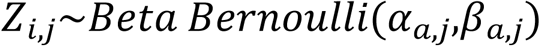

Where α_*a*,*j*_ and β_*a*,*j*_are the shape and scale parameters of the Beta distribution. We assumed α = 1 and β = 10 for all times (*j*) and all age groups (*a*), which corresponds to a prior mean probability of infection of 0.1 and prior variance of 0.0826 in each norovirus season.

#### Antibody Kinetics Model

Individuals have unique infection histories, but the parameters governing post-infection antibody kinetics were assumed to be universal. An individual’s predicted antibody level at time *t* against variant *j* is given by:

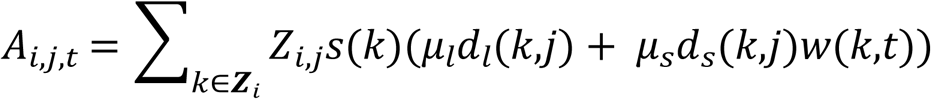

Where *k* represents the *k*th variant in the individual’s infection history, *s*(*k*) represents the degree of immune suppression on the overall boost from previous exposures, μ_*l*_ represents the level of long-term homologous boosting, μ_*s*_ represents the level of transient homologous boosting, *d*_*l*_(*k*,*j*) represents the level of long-term cross-reactivity between infecting variant *k* and measured variant *j* (governed by the antigenic map), *d*_*s*_(*k*,*j*) represents the level of transient cross-reactivity, and *w*(*k*,*t*) represents the level of waning of the transient response from infection with variant *k* by observation time *t*.

We ran two versions of the serosolver antibody kinetics model. The first used the published version of serosolver, where antibody levels from the short-term, transient boost are assumed to decline linearly on the log-scale with time-since-infection. We refer to this as the “linear” model:

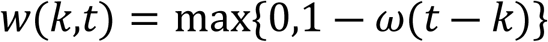

Where ω is the waning rate parameter. Similarly, this version of the model assumes that antibody boosting declines linearly as a function of antigenic distance between the infecting strain and the measured strain:

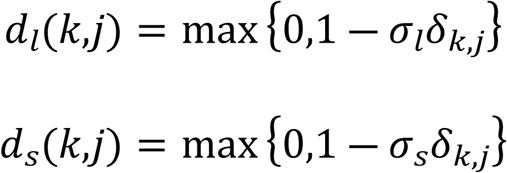

Where σ_*l*_ and σ_*s*_ are the cross-reactivity gradient parameters for the long and short-term response and δ_*k*,*j*_ is the Euclidean distance between variant *k* and variant *j* on the antigenic map.

Second, we implemented an alternative antibody kinetics model which replaced these linear decay models with exponential decay models (again on the log-scale):

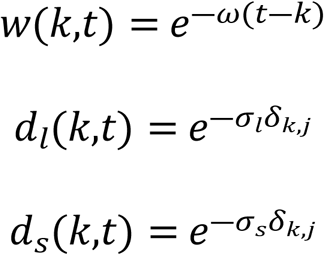

We refer to this as the “exponential” model. The number of parameters in the exponential model are the same as in the linear model, and the interpretation of all other parameters is unchanged.

Immune suppression with each successive boost was modelled as:

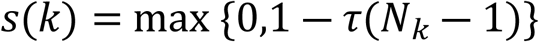

Where τ is the immune suppression parameter, and *N*_*k*_ denotes that the infection with variant *k* is the *N*_*k*_th infection in the individual’s infection history.

#### Observation Model

The original version of serosolver assumes normally distributed observations on the log scale with censoring to account for the upper and lower bounds of the assay. This is the version we use for the linear antibody kinetics model. However, we noticed that this model underestimated the number of negative (undetectable) observations. Therefore, we updated the observation model for the exponential model to include an additional spike-and-slab distribution. Where the model-predicted antibody levels are negative (i.e., where an individual has not been previously infected), observed titres are assumed to be distributed according to the mixture of a spike and slab distribution: with probability 0.001, negative antibody levels are uniformly distributed within the bounds of the assay, and otherwise indetectable with probability 0.999. Model-predicted positive log antibody levels are assumed to be normally distributed as described in the original observation model.

#### Model Fitting

Parameters were estimated using a custom adaptive Metropolis-Hastings algorithm, full details given in (23,47). For the model results presented, 5 chains were run for 1,000,000 iterations, with the first 300,000 iterations discarded, and thinned to every 500^th^ iteration, returning 2,000 samples of the posterior. Parameter priors are given in Appendix, Table A3 and prior predictive plots illustrated in Fig. A9. Convergence was assessed based on effective sample sizes exceeding 200 for all parameters and the upper 95% CI of the Gelman-Rubin statistic being below 1.1 Example traceplots of the posterior distributions are given in the appendix (Appendix Fig. A10), and density plots of the priors and posterior estimates are also provided (Appendix, Fig. A11). A comparison of the ic_50_ antibody data of each norovirus variant and the estimated probability of infection in a given time window is illustrated in Appendix, Fig. A12 for a sample of individuals. A good fit to the data should correspond with a raised antibody titre to a specific infection corresponding with a high probability of infection. This is apparent in sample individuals 2, 5, 7, 19, but is it also apparent that a number of individuals have no response to any variants, likely corresponding to non-responders. Posterior estimates of the antibody kinetics model assuming a single infection (at time t=0) is illustrated in Appendix Fig. 13, illustrating the inferred homotypic and heterotypic responses.

Because the age distribution of our sample did not reflect the age distribution of the English under 5 population (Appendix Fig. A2), we adjusted our attack rate estimates by resampling individuals with replacement for each posterior draw until the distribution of ages in the sample matched the age distribution from ONS census data (https://www.ons.gov.uk/peoplepopulationandcommunity/populationandmigration/populationestimates/datasets/populationestimatesforukenglandandwalesscotlandandnorthernireland).

The data and code for the analysis are available here: https://github.com/kath-o-reilly/serosolver-norovirus-eng-serology-v2

## Data Availability

Both the data and code used to run the analyses are available in publicly accessible repositories

https://zenodo.org/records/7547170

https://github.com/kath-o-reilly/serosolver-norovirus-eng-serology-v2

## Acknowledgments

Funding was provided by the Wellcome Trust [203268/Z/16/Z] and the National Institute for Health and Care Research (NIHR) Health Protection Research Unit in Modelling and Health Economics, a partnership between the UK Health Security Agency, Imperial College London and LSHTM (grant code NIHR200908). JAH is supported by a Wellcome Trust Early Career Award (grant 225001/Z/22/Z). We acknowledge input given by the Enteric Virus Unit at UKHSA, and provision of data describing annual proportions of norovirus GII.4 variants.

Preliminary results of this analysis were presented at the 2023 Calicivirus Conference in Rotterdam, Netherlands, and we acknowledge the feedback received.

## References

1. Bartel B, Gau E. Nosocomial diarrhea: a review of pathophysiology, etiology, and treatment strategies. Hosp Pract 1995. 2012 Feb;40(1):130–8. doi:10.3810/hp.2012.02.953 PubMed PMID: 22406888.

2. Galles NC, Liu PY, Updike RL, Fullman N, Nguyen J, Rolfe S, et al. Measuring routine childhood vaccination coverage in 204 countries and territories, 1980–2019: a systematic analysis for the Global Burden of Disease Study 2020, Release 1. The Lancet. 2021 Aug 7;398(10299):503–21. doi:10.1016/S0140-6736(21)00984-3 PubMed PMID: 34273291.

3. Nielsen J, Vestergaard LS, Richter L, Schmid D, Bustos N, Asikainen T, et al. European all-cause excess and influenza-attributable mortality in the 2017/18 season: should the burden of influenza B be reconsidered? Clin Microbiol Infect Off Publ Eur Soc Clin Microbiol Infect Dis. 2019 Oct;25(10):1266–76. doi:10.1016/j.cmi.2019.02.011 PubMed PMID: 30790685.

4. Beek J van, Graaf M de, Al-Hello H, Allen DJ, Ambert-Balay K, Botteldoorn N, et al. Molecular surveillance of norovirus, 2005–16: an epidemiological analysis of data collected from the NoroNet network. Lancet Infect Dis. 2018 May 1;18(5):545–53. doi:10.1016/S1473-3099(18)30059-8 PubMed PMID: 29396001.

5. Lindesmith LC, Boshier FAT, Brewer-Jensen PD, Roy S, Costantini V, Mallory ML, et al. Immune Imprinting Drives Human Norovirus Potential for Global Spread. mBio. 2022 Sep 14;13(5):e01861–22. doi:10.1128/mbio.01861-22

6. Beek J van, Ambert-Balay K, Botteldoorn N, Eden JS, Fonager J, Hewitt J, et al. Indications for worldwide increased norovirus activity associated with emergence of a new variant of genotype II.4, late 2012. Eurosurveillance. 2013 Jan 3;18(1):20345. doi:10.2807/ese.18.01.20345-en

7. Kroneman A, Vennema H, Duijnhoven Y van, Duizer E, Koopmans M. High number of norovirus outbreaks associated with a GGII.4 variant in the Netherlands and elsewhere: does this herald a worldwide increase? Wkly Releases 1997–2007. 2004 Dec 23;8(52):2606. doi:10.2807/esw.08.52.02606-en

8. Lopman BA, Adak GK, Reacher MH, Brown DWG. Two epidemiologic patterns of norovirus outbreaks: surveillance in England and wales, 1992-2000. Emerg Infect Dis. 2003 Jan;9(1):71–7. doi:10.3201/eid0901.020175 PubMed PMID: 12533284; PubMed Central PMCID: PMC2873766.

9. Lopman B, Vennema H, Kohli E, Pothier P, Sanchez A, Negredo A, et al. Increase in viral gastroenteritis outbreaks in Europe and epidemic spread of new norovirus variant. The Lancet. 2004 Feb 28;363(9410):682–8. doi:10.1016/S0140-6736(04)15641-9

10. Rouzine IM, Rozhnova G. Antigenic evolution of viruses in host populations. PLoS Pathog. 2018 Sep;14(9):e1007291. doi:10.1371/journal.ppat.1007291

11. Becker M, Strengert M, Junker D, Kaiser PD, Kerrinnes T, Traenkle B, et al. Exploring beyond clinical routine SARS-CoV-2 serology using MultiCoV-Ab to evaluate endemic coronavirus cross-reactivity. Nat Commun. 2021 Feb 19;12(1):1152. doi:10.1038/s41467-021-20973-3

12. Reeck A, Kavanagh O, Estes MK, Opekun AR, Gilger MA, Graham DY, et al. Serological correlate of protection against norovirus-induced gastroenteritis. J Infect Dis. 2010 Oct 15;202(8):1212–8. doi:10.1086/656364 PubMed PMID: 20815703; PubMed Central PMCID: PMC2945238.

13. Lindesmith LC, Donaldson EF, Baric RS. Norovirus GII.4 strain antigenic variation. J Virol. 2011 Jan;85(1):231–42. doi:10.1128/JVI.01364-10 PubMed PMID: 20980508; PubMed Central PMCID: PMC3014165.

14. Harrington PR, Lindesmith L, Yount B, Moe CL, Baric RS. Binding of Norwalk virus-like particles to ABH histo-blood group antigens is blocked by antisera from infected human volunteers or experimentally vaccinated mice. J Virol. 2002 Dec;76(23):12335–43. doi:10.1128/jvi.76.23.12335-12343.2002 PubMed PMID: 12414974; PubMed Central PMCID: PMC136916.

15. Debbink K, Lindesmith LC, Donaldson EF, Costantini V, Beltramello M, Corti D, et al. Emergence of New Pandemic GII.4 Sydney Norovirus Strain Correlates With Escape From Herd Immunity. J Infect Dis. 2013 Dec 1;208(11):1877–87. doi:10.1093/infdis/jit370

16. Kendra JA, Tohma K, Ford-Siltz LA, Lepore CJ, Parra GI. Antigenic cartography reveals complexities of genetic determinants that lead to antigenic differences among pandemic GII.4 noroviruses. Proc Natl Acad Sci. 2021 Mar 16;118(11):e2015874118. doi:10.1073/pnas.2015874118

17. Debbink K, Lindesmith LC, Ferris MT, Swanstrom J, Beltramello M, Corti D, et al. Within-host evolution results in antigenically distinct GII.4 noroviruses. J Virol. 2014 Jul;88(13):7244–55. doi:10.1128/JVI.00203-14 PubMed PMID: 24648459; PubMed Central PMCID: PMC4054459.

18. Debbink K, Lindesmith LC, Donaldson EF, Swanstrom J, Baric RS. Chimeric GII.4 Norovirus Virus-Like-Particle-Based Vaccines Induce Broadly Blocking Immune Responses. J Virol. 2014 Jul;88(13):7256–66. doi:10.1128/jvi.00785-14

19. Oidtman RJ, Arevalo P, Bi Q, McGough L, Russo CJ, Vera Cruz D, et al. Influenza immune escape under heterogeneous host immune histories. Trends Microbiol. 2021 Dec 1;29(12):1072–82. doi:10.1016/j.tim.2021.05.009

20. Lindesmith LC, Ferris MT, Mullan CW, Ferreira J, Debbink K, Swanstrom J, et al. Broad Blockade Antibody Responses in Human Volunteers after Immunization with a Multivalent Norovirus VLP Candidate Vaccine: Immunological Analyses from a Phase I Clinical Trial. PLoS Med. 2015 Mar;12(3):e1001807. doi:10.1371/journal.pmed.1001807

21. Lindesmith LC, Brewer-Jensen PD, Conrad H, O’Reilly KM, Mallory ML, Kelly D, et al. Emergent variant modeling of the serological repertoire to norovirus in young children. Cell Rep Med. 2023 Mar 21;4(3):100954. doi:10.1016/j.xcrm.2023.100954

22. Smith DJ, Lapedes AS, de Jong JC, Bestebroer TM, Rimmelzwaan GF, Osterhaus ADME, et al. Mapping the antigenic and genetic evolution of influenza virus. Science. 2004 Jul 16;305(5682):371–6. doi:10.1126/science.1097211 PubMed PMID: 15218094.

23. Hay JA, Minter A, Ainslie KEC, Lessler J, Yang B, Cummings DAT, et al. An open source tool to infer epidemiological and immunological dynamics from serological data: serosolver. PLoS Comput Biol. 2020 May;16(5):e1007840. doi:10.1371/journal.pcbi.1007840 PubMed PMID: 32365062; PubMed Central PMCID: PMC7241836.

24. Fonville JM, Wilks SH, James SL, Fox A, Ventresca M, Aban M, et al. Antibody landscapes after influenza virus infection or vaccination. Science. 2014 Nov 21;346(6212):996–1000. doi:10.1126/science.1256427 PubMed PMID: 25414313; PubMed Central PMCID: PMC4246172.

25. Quee FA, de Hoog MLA, Schuurman R, Bruijning-Verhagen P. Community burden and transmission of acute gastroenteritis caused by norovirus and rotavirus in the Netherlands (RotaFam): a prospective household-based cohort study. Lancet Infect Dis. 2020 May 1;20(5):598–606. doi:10.1016/S1473-3099(20)30058-X

26. O’Brien SJ, Donaldson AL, Iturriza-Gomara M, Tam CC. Age-Specific Incidence Rates for Norovirus in the Community and Presenting to Primary Healthcare Facilities in the United Kingdom. J Infect Dis. 2016 Feb 1;213 Suppl 1:S15–18. doi:10.1093/infdis/jiv411 PubMed PMID: 26744427; PubMed Central PMCID: PMC4704656.

27. El-Heneidy A, Grimwood K, Mihala G, Lambert S, Ware RS. Epidemiology of Norovirus in the First 2 Years of Life in an Australian Community-based Birth Cohort. Pediatr Infect Dis J. 2022 Nov;41(11):878. doi:10.1097/INF.0000000000003667

28. Measuring social networks in British primary schools through scientific engagement [Internet]. [cited 2025 Nov 27]. Available from: https://royalsocietypublishing.org/doi/epdf/10.1098/rspb.2010.1807 doi:10.1098/rspb.2010.1807

29. Lindesmith LC, Donaldson EF, Lobue AD, Cannon JL, Zheng DP, Vinje J, et al. Mechanisms of GII.4 norovirus persistence in human populations. Plos Med. 2008 Feb;5(2):269–90. doi:10.1371/journal.pmed.0050031

30. GOV.UK [Internet]. [cited 2026 Feb 23]. Norovirus: annual figures 2000 to 2012. Available from: https://www.gov.uk/government/publications/norovirus-annual-figures-2000-to-2012

31. Lindesmith LC, McDaniel JR, Changela A, Verardi R, Kerr SA, Costantini V, et al. Sera Antibody Repertoire Analyses Reveal Mechanisms of Broad and Pandemic Strain Neutralizing Responses after Human Norovirus Vaccination. Immunity. 2019 Jun 18;50(6):1530–1541.e8. doi:10.1016/j.immuni.2019.05.007 PubMed PMID: 31216462; PubMed Central PMCID: PMC6591005.

32. Lindesmith LC, Brewer-Jensen PD, Mallory ML, Zweigart MR, May SR, Kelly D, et al. Antigenic Site Immunodominance Redirection Following Repeat Variant Exposure. Viruses. 2022 Jun;14(6):6. doi:10.3390/v14061293

33. Lessler J, Riley S, Read JM, Wang S, Zhu H, Smith GJD, et al. Evidence for Antigenic Seniority in Influenza A (H3N2) Antibody Responses in Southern China. PLoS Pathog. 2012 Jul;8(7):e1002802. doi:10.1371/journal.ppat.1002802

34. Voss WN, Mallory MA, Byrne PO, Marchioni JM, Knudson SA, Powers JM, et al. Hybrid immunity to SARS-CoV-2 arises from serological recall of IgG antibodies distinctly imprinted by infection or vaccination. Cell Rep Med. 2024 Aug 20;5(8):101668. doi:10.1016/j.xcrm.2024.101668 PubMed PMID: 39094579; PubMed Central PMCID: PMC11384961.

35. Gaythorpe KAM, Trotter CL, Conlan AJK. Modelling norovirus transmission and vaccination. Vaccine. 2018 Sep 5;36(37):5565–71. doi:10.1016/j.vaccine.2018.07.053

36. Vesga JF, Douglas A, Celma C, Knock ES, Baguelin M, Edmunds WJ. The transmission dynamics of Norovirus in England: A genotype-specific modelling study. Epidemics. 2025 Nov 27;53:100875. doi:10.1016/j.epidem.2025.100875 PubMed PMID: 41319617.

37. Waerlop G, Janssens Y, Jacobs B, Jarczowski F, Diessner A, Leroux-Roels G, et al. Immune responses in healthy adults elicited by a bivalent norovirus vaccine candidate composed of GI.4 and GII.4 VLPs without adjuvant. Front Immunol. 2023;14:1188431. doi:10.3389/fimmu.2023.1188431 PubMed PMID: 37435073; PubMed Central PMCID: PMC10331465.

38. Ghosh S, Malik YS, Kobayashi N. Therapeutics and Immunoprophylaxis Against Noroviruses and Rotaviruses: The Past, Present, and Future. Curr Drug Metab. 2018;19(3):170–91. doi:10.2174/1389200218666170912161449 PubMed PMID: 28901254; PubMed Central PMCID: PMC5971199.

39. Chhabra P, de Graaf M, Parra GI, Chan MCW, Green K, Martella V, et al. Updated classification of norovirus genogroups and genotypes. J Gen Virol. 2019 Oct;100(10):1393–406. doi:10.1099/jgv.0.001318 PubMed PMID: 31483239; PubMed Central PMCID: PMC7011714.

40. Allen DJ, Trainor E, Callaghan A, O’Brien SJ, Cunliffe NA, Iturriza-Gómara M. Early Detection of Epidemic GII-4 Norovirus Strains in UK and Malawi: Role of Surveillance of Sporadic Acute Gastroenteritis in Anticipating Global Epidemics. PLOS ONE. 2016 Apr 26;11(4):e0146972. doi:10.1371/journal.pone.0146972

41. Chhabra P, Wong S, Niendorf S, Lederer I, Vennema H, Faber M, et al. Increased circulation of GII.17 noroviruses, six European countries and the United States, 2023 to 2024. Eurosurveillance. 2024 Sep 26;29(39):2400625. doi:10.2807/1560-7917.ES.2024.29.39.2400625

42. Villabruna N, Izquierdo-Lara RW, Schapendonk CME, de Bruin E, Chandler F, Thao TTN, et al. Profiling of humoral immune responses to norovirus in children across Europe. Sci Rep. 2022 Aug 22;12(1):14275. doi:10.1038/s41598-022-18383-6 PubMed PMID: 35995986; PubMed Central PMCID: PMC9395339.

43. Allen DJ, Adams NL, Aladin F, Harris JP, Brown DWG. Emergence of the GII-4 Norovirus Sydney2012 strain in England, winter 2012-2013. PloS One. 2014;9(2):e88978. doi:10.1371/journal.pone.0088978 PubMed PMID: 24551201; PubMed Central PMCID: PMC3923861.

44. Atmar RL, Ettayebi K, Ayyar BV, Neill FH, Braun RP, Ramani S, et al. Comparison of Microneutralization and Histo-Blood Group Antigen-Blocking Assays for Functional Norovirus Antibody Detection. J Infect Dis. 2020 Feb 18;221(5):739–43. doi:10.1093/infdis/jiz526 PubMed PMID: 31613328; PubMed Central PMCID: PMC8483564.

45. Koromyslova AD, Morozov VA, Hefele L, Hansman GS. Human Norovirus Neutralized by a Monoclonal Antibody Targeting the Histo-Blood Group Antigen Pocket. J Virol. 2019 Mar 1;93(5):e02174–18. doi:10.1128/JVI.02174-18 PubMed PMID: 30541855; PubMed Central PMCID: PMC6384083.

46. Alvarado G, Ettayebi K, Atmar RL, Bombardi RG, Kose N, Estes MK, et al. Human Monoclonal Antibodies That Neutralize Pandemic GII.4 Noroviruses. Gastroenterology. 2018 Dec;155(6):1898–907. doi:10.1053/j.gastro.2018.08.039 PubMed PMID: 30170116; PubMed Central PMCID: PMC6402321.

47. Hay JA, Zhu H, Jiang CQ, Kwok KO, Shen R, Kucharski A, et al. Reconstructed influenza A/H3N2 infection histories reveal variation in incidence and antibody dynamics over the life course. PLOS Biol. 2024 Nov 7;22(11):e3002864. doi:10.1371/journal.pbio.3002864

